# Quantifying the Optimism of Naive Cross-Validation for Binary Outcome Prediction with Repeated-Measures Predictors: A Simulation Study and Clinical Illustration

**DOI:** 10.64898/2026.05.27.26354222

**Authors:** Joseph L. Hagan

## Abstract

**Background.:** Cross-validation (CV) is widely used to estimate the discriminative performance of clinical prediction models, but applied at the observation level to repeated-measures data it can overestimate performance. A common but under-characterized structure in neonatal, critical care, and wearable-sensor research pairs continuous predictors measured repeatedly within subjects with a binary subject-level event, evaluated by area under the receiver operating characteristic curve (AUROC). The magnitude of this optimism remains uncharacterized against true out-of-sample performance.

**Methods.:** Three strategies for estimating subject-level AUROC from ridge logistic regression were compared: naive observation-level 10-fold CV, subject-level 10-fold CV, and leave-one-cluster-out (LOCO) CV. They were applied to daily oxygenation measures and retinopathy of prematurity (ROP) outcomes in 101 extremely low birth weight infants, and to a factorial simulation of 162 conditions varying cluster count (20 to 150), intraclass correlation (ICC, 0.1 to 0.5), lag-1 autocorrelation (0.2 to 0.8), and prevalence (10% to 35%). An independent sample provided a true-performance reference, decomposing optimism into a partitioning component and a residual discrepancy. A paired 24-condition arm held latent signal variance constant across ICC.

**Results.:** In the clinical dataset, naive CV optimism was +0.078 AUROC units for severe ROP and +0.031 for any ROP. In the simulation, total optimism relative to true performance averaged +0.213, comprising a partitioning component (mean +0.154, range +0.071 to +0.257) and a residual internal-validation discrepancy (mean +0.059). The partitioning component increased with higher ICC and autocorrelation, fewer clusters, and lower event rates, whereas the residual discrepancy did not rise with dependence. Under signal control, the rise in the partitioning component across ICC was largely retained (+0.041 versus +0.052) whereas the decline in the residual was attenuated by 73%. Subject-level 10-fold CV closely agreed with LOCO throughout (mean absolute deviation 0.004 in simulation).

**Conclusions.:** Naive observation-level CV meaningfully overestimates discriminative performance; subject-level partitioning eliminates the partitioning component and closely agrees with LOCO, although a residual discrepancy from true performance remains at small cluster counts. Subject-level partitioning should be considered essential rather than optional, at every level of nested resampling.

## 1. Background

Clinical prediction models for binary outcomes support risk stratification and therapeutic decision-making across medicine. The area under the receiver operating characteristic curve (AUROC) is the most widely reported measure of discriminative performance, providing a threshold-independent summary of the probability that a randomly selected case receives a higher predicted risk than a randomly selected non-case [1]. Reliable estimation of AUROC is therefore central to the evaluation of any candidate model.

Cross-validation (CV) is among the most commonly used internal validation strategies, providing resampling-based performance estimation without a dedicated holdout dataset [2,3]. Established approaches, including k-fold CV, leave-one-out CV, and bootstrap-based optimism correction, offer different tradeoffs between bias, variance, and computation [4,5]. These methods share a common assumption: that individual observations are exchangeable, so that any partition into training and test sets yields conditionally independent subsets. When this assumption holds, k-fold CV provides approximately unbiased estimates of out-of-sample performance [3]. When it is violated, CV-based estimates can be substantially optimistic, particularly when the same subjects contribute observations to both training and test folds [6].

In many clinical settings this assumption is systematically violated. Longitudinal studies, electronic health record cohorts, continuous or high-frequency physiological monitoring, and other repeated-measures data sources generate multiple observations per subject over time, inducing within-subject serial dependence. When naive CV partitions data at the observation level, assigning individual time points to folds without regard to cluster membership, training and test sets routinely contain observations from the same subject. This within-subject contamination produces optimistically biased performance estimates, because held-out observations from previously seen subjects are easier to predict than observations from entirely new subjects. Leakage of this general kind has been identified as a recurrent source of irreproducible performance claims across machine-learning-based science [7]. Subject-wise partitioning is required to obtain estimates that generalize to previously unseen subjects.

### Sources of optimism in naive CV

Naive observation-level CV in this setting generates optimism through two qualitative pathways. The first is direct leakage through within-subject temporal dependence: a predictor trajectory partially observed during training carries information about the same subject’s unobserved observations in the test set, making held-out predictions artificially accurate. The second is indirect leakage through the outcome: because the binary outcome is constant within each subject, the presence of any observation from a subject in the training set conveys information about that subject’s remaining observations. These two pathways operate through the within-subject temporal component (autocorrelation) and the between-subject component (ICC), respectively, so that both sources of dependence contribute to optimism. The present study treats these pathways qualitatively. Their formal separation within this covariance framework is developed in Appendix C. A fuller formal and empirical treatment, including fixed baseline covariates as the limiting case of time-invariant predictors, is the subject of a manuscript in preparation.

### Relationship to existing work

Inter-cluster contamination in CV has been recognized across domains. Roberts et al. [8] showed that naive random CV overestimates transferability under spatial autocorrelation and proposed block partitioning; Bergmeir and colleagues [9,10] showed that observation-level splits yield optimistic error estimates for autoregressive time series. A recent overview of generalization-error estimation across nonstandard data settings reached a convergent conclusion for clustered data, recommending grouped partitioning, but explicitly excluded repeated measurements over time from its treatment [11]. The closest methodological precedent for the present structure is Saeb et al. [12], who simulated the repeated-measures predictor, subject-level binary outcome setting while varying between-subject variance, within-subject variance, and subject count, and who reported that approximately 45% of surveyed studies using repeated sensor measurements employed naive CV [12]. That work fixed prevalence at 50%, reported random-forest classification error rather than AUROC, and generated within-subject observations as independent draws given fixed subject-level effects, so that temporal autocorrelation was absent from the data-generating model. A separate and rapidly developing line of work has formalized CV for correlated data: Rabinowicz and Rosset [13] derived the bias of standard CV under within-cluster correlation and a covariance-corrected estimator, and Yuval and Rosset [14] extended this treatment to classification. That work corrects a CV performance estimate given a specified covariance model, whereas the present study requires no such specification, instead providing an interpretable, factorial quantification of naive-CV optimism as a function of clinically measurable parameters (cluster count, ICC, autocorrelation, prevalence) for the subject-level binary outcome, a direct comparison of naive against cluster-aware partitioning, and a decomposition of the resulting optimism against true out-of-sample performance. Together with the exclusion of within-subject repeated measures from TRIPOD-Cluster [15], the multicenter-focused extension of the original TRIPOD statement [16], this indicates that the commonly encountered structure addressed here falls outside existing general clustered-data and multicenter validation guidance, consistent with a recent broad appraisal of clinical prediction modeling that identifies heterogeneity across clusters and inadequate validation methodology as persistent, unresolved challenges for the field [17].

The methodological contribution of this study is the first factorial quantification of naive- CV optimism against a genuine out-of-sample reference for the repeated-measures predictor, subject-level binary outcome data structure, together with its decomposition into a leakage-driven partitioning component and a sample-size-governed residual internal-validation discrepancy that respond differently to within-subject dependence, and actionable guidance for the applied literature. The advance is therefore one of quantification and decomposition rather than the introduction of a new algorithm.

## 2. Methods

### 2.1 Motivating dataset

The framework was illustrated using data from a previously published study of oxygenation factors and retinopathy of prematurity (ROP) among extremely low birth weight infants [18], in which the present author served as co-investigator and biostatistician. That study examined oxygenation characteristics during the first two postnatal months among infants with birth weight below 1000 grams admitted to a level III neonatal intensive care unit (NICU) at Northside Hospital in Atlanta, Georgia, born between January 1, 2016, and December 31, 2020. Data comprised daily oxygenation variables derived from continuous pulse oximetry (SpO ) and inspired oxygen (FiO ) recordings. The study was conducted under a de-identified data protocol approved by the Northside Hospital Institutional Review Board, and no identifying information was used in the present secondary analysis. The motivating dataset was used solely as aclinical illustration of validation bias and was not intended to develop a deployable ROP prediction model.

The population comprised 101 infants meeting inclusion criteria, including availability of simultaneous SpO and FiO data for at least 45 of the first 60 days of life and survival to eye examination. In this structure, each infant constitutes a cluster, the 101 infants comprise the cluster count, and the repeated daily observations serve as the within-cluster units. Each infant contributed between 47 and 60 daily observations (median = 59), yielding 5,753 observation-level rows; after exclusion of 138 observations with missing FiO -dependent variables (2.4%), the complete-case dataset comprised 5,615 observations from all 101 subjects, with 44 to 60 observations per infant (median = 57). ROP was classified as no ROP (n = 53), mild ROP (stages 1 to 2, n = 33), and severe ROP (stage 3, n = 15). For the primary analysis, ROP was dichotomized as severe versus not severe (15 events; prevalence 14.9%). For the secondary analysis, ROP was dichotomized as any versus none (48 events; prevalence 47.5%). The severe ROP dichotomization was selected as primary because it represents the clinically actionable outcome and because its low event count places it in the small-sample regime most vulnerable to CV-related bias. Throughout, the terms *subject* and *cluster* are used synonymously for the unit of clustering.

### 2.2 Predictor selection

The original dataset provided 12 daily oxygenation variables per subject exhibiting extensive multicollinearity (maximum pairwise |r| = 0.96). The predictor set was reduced to seven oxygenation variables plus day of life (DOL), selected on clinical grounds across five physiological domains. For oxygen delivery, mean daily FiO (Avg_FiO ), significantly elevated in severe ROP in the original study (adjusted P = .001), and the titration index (Tit_Index), a distinct measure of manual FiO adjustment effort, were retained. For overall saturation, mean daily SpO (Avg_SpO ) was retained, significantly lower in severe ROP in the original study (adjusted P < .001) and correlated above |r| = 0.7 with the hypoxemia index (r = −0.96), hypoxemic episodes (r = −0.77), and ambient hyperoxemia (r = +0.71), so that these near-redundant variables were excluded. For hypoxemia severity, the percentage of time in severe hypoxemia (SpO < 80%) was retained, showing the strongest hypoxemia association with severe ROP (adjusted P = .005). For hyperoxemia subtype, ambient air hyperoxemia (more frequent without ROP, adjusted P = .003) and iatrogenic hyperoxemia (associated with severe ROP, adjusted P = .046) were both retained because of their opposing associations, and the composite hyperoxemia index that conflates them was excluded. For oxygenation lability, the daily count of rapid oscillations between hypoxemia and hyperoxemia (Swings) captured a construct not represented by any level or duration variable. After trimming, the maximum pairwise correlation among the seven retained oxygenation predictors was |r| = 0.71 (Avg_SpO and ambient hyperoxemia; Appendix A), reduced from |r| = 0.96.

### 2.3 Within-cluster dependence structure

Two complementary measures were estimated for each of the seven oxygenation predictors. The intraclass correlation coefficient (ICC), in the one-way random-effects form (ICC(1,1) of Shrout and Fleiss [19]), was estimated as ICC = (MS_between − MS_within) / (MS_between + (n − 1) × MS_within), where MS_between and MS_within denote the between-cluster and within-cluster mean squares and n the average cluster size. Because cluster sizes were nearly balanced, the arithmetic mean cluster size (55.59) was used for n ; substituting the effective cluster size for an unbalanced one-way design, (N − Σn ² / N) / (k − 1) = 55.59, where N denotes the total number of observations and k the number of clusters, altered every reported ICC estimate by less than 0.0001. Observations were ordered by DOL within each infant before the lag-1 autocorrelations were computed. The ICC quantifies the proportion of total variance attributable to stable between-subject differences and is a single value per predictor across all subjects; it is distinct from the within-subject lag-1 autocorrelation, which quantifies day-to-day serial dependence in a predictor’s within-subject deviations. ICC values ranged from 0.16 (iatrogenic hyperoxemia) to 0.53 (Avg_FiO ), and median within-cluster lag-1 autocorrelations ranged from 0.45 to 0.73 (Appendix B). This combination of moderate-to-high ICC and substantial autocorrelation is precisely the condition under which naive observation-level CV is expected to be optimistically biased.

### 2.4 Prediction model

All models were fit using ridge (L2-penalized) logistic regression, with the linear predictor comprising DOL and the seven oxygenation variables of Section 2.2. Ridge penalization was applied to all coefficients except the intercept, with a fixed regularization parameter (λ = 0.01). Class weights were balanced inversely to class frequency to accommodate the low event rate for severe ROP, and all predictors were standardized to zero mean and unit variance within training folds, using the internal standardization of the fitting routine computed on the class-weighted training sample. Ridge regression was selected because the moderate collinearity among oxygenation predictors (maximum |r| = 0.71) and the extreme events-per-variable ratio (as few as 15 events with 8 predictors) required regularization. A fixed λ was preferred over fold-specific tuning to avoid introducing variability into the AUROC estimates that could not be cleanly attributed to the partitioning strategy; a fixed value of 0.01 matched the simulation design, and a regularization sensitivity analysis (Section 3.5) confirmed that the optimism findings were robust across a 100,000-fold range of λ.

### 2.5 Cross-validation strategies

Three CV strategies were compared, each applied to the same ridge model. For every strategy, AUROC was computed at the subject level: each subject’s held-out daily predicted probabilities were averaged to a single subject-level prediction, and AUROC was calculated over these subject-level predictions. Averaging aligns the prediction with the subject-level outcome and ensures that each subject contributes one prediction to the AUROC without additional modeling assumptions about the temporal trajectory. The three partitioning strategies (Figure 1) were: *naive observation-level 10-fold CV*, in which daily observations were randomly assigned to 10 folds without regard to subject membership (the methodologically incorrect approach); *subject-level 10-fold CV*, in which subjects were assigned to 10 folds so that all of a subject’s observations shared a fold, eliminating within-subject leakage; and *leave-one-cluster-out (LOCO) CV*, in which each subject was held out in turn with the model trained on the remaining subjects.

**Figure 1.**
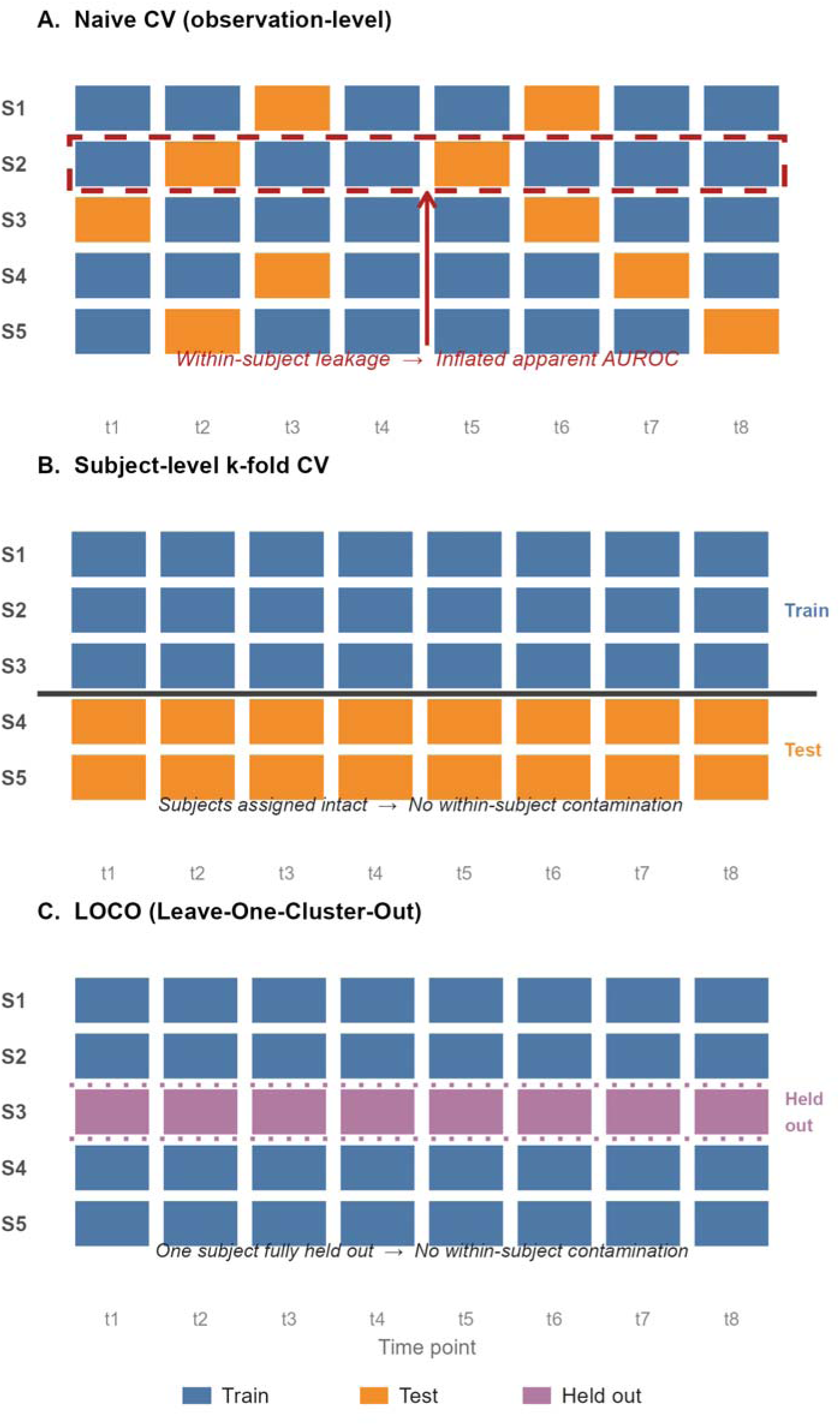
Illustration of cross-validation partitioning strategies for repeated-measures data with subject-level outcomes. Each row represents one subject, and each cell represents one observation at a time point. Panel A depicts naive observation-level CV, in which observations are assigned to folds without regard to subject membership, illustrating within-subject leakage. Panel B depicts subject-level k-fold CV, in which all of a subject’s observations are assigned intact to one fold, so that the test fold contains complete subjects rather than individual observations. Panel C depicts leave-one- cluster-out CV, in which one subject is fully withheld as the test case at each iteration (one representative iteration is shown), with this role rotating across all subjects in turn.

Under LOCO, each held-out subject yields exactly one out-of-sample prediction, and these predictions are pooled across all N iterations into a single AUROC computed over the N subjects; LOCO was treated as a leakage-free reference because each held-out subject’s observations are entirely absent from training and because its deterministic structure removes fold-assignment variability.

For the motivating dataset, two subject-level summary approaches were additionally evaluated, in which daily series were reduced to per-subject statistics (means only, 8 features; or means plus linear slopes over DOL, 15 features) and logistic regression was fit on the resulting 101-row dataset.

### 2.6 Evaluation framework

The primary quantity was optimism. For the simulation, optimism was decomposed by reference to true out-of-sample performance (Section 2.7): the *partitioning component* was defined as the difference between naive CV AUROC and subject-level CV AUROC (the leakage that cluster-aware partitioning removes), and the *residual internal-validation discrepancy* as the difference between subject-level CV AUROC and the true AUROC of the full-data model on the independent test sample. This residual is a descriptive component of the additive decomposition, not a pure estimate of finite-sample optimism, because the CV models and the full-data model are trained on different numbers of subjects; within the conditions examined, it was governed primarily by development-sample size and was not materially affected by the choice of k for subject-level CV. For the motivating dataset, where no population is available to sample, optimism was defined relative to LOCO (AUROC_CV − AUROC_LOCO); because subject-level CV and LOCO coincide in the simulation, this LOCO-referenced quantity estimates the partitioning component. For the LOCO AUROC, a subject-level (cluster) bootstrap 95% confidence interval was obtained by resampling subjects with replacement and recomputing the pooled subject-level AUROC (2,000 resamples), which does not rely on the single-fixed-model assumption underlying the DeLong method [1] and is therefore appropriate when each held-out prediction comes from a different model. For the k-fold strategies, replicate random fold assignments (50 for the motivating dataset, 10 in the simulation) characterized partitioning-related variability; the ratio of naive to subject-level CV standard deviation quantified the artificial precision ("precision illusion") produced by naive partitioning. These replicate-based intervals and standard deviations characterize variability with respect to random fold assignment only and should not be interpreted as confidence intervals or standard errors for the population AUROC. AUROC was the sole metric, reflecting the study objective of quantifying discrimination bias; calibration was not evaluated.

### 2.7 Simulation study

#### Data-generating mechanism

For each cluster i (i = 1, …, N), a vector of cluster-level random effects b_i was drawn from a multivariate normal distribution with a mild factor structure inducing between-predictor correlation. For each time point t (t = 1, …, 57), predictor values were generated as x_it = b_i + e_it, where the within-cluster errors followed a first-order autoregressive AR(1) process e_it = ρ e_{i,t−1} + ε_it, with innovation variance σ²_ε = σ²_e(1 − ρ²), σ²_e = 1 − ICC, and between-cluster variance σ²_b = ICC, so that each predictor had total marginal variance 1.0. The within-subject innovations ε_it were drawn independently across predictors at each time point; cross-predictor dependence entered only through the between-subject random effects b_i and their correlation matrix R (below). A single unit-diagonal predictor correlation matrix R (two latent factors, loadings drawn independently from N(0, 0.30²), unique variance 0.70 per predictor; maximum absolute off-diagonal correlation, 0.211) was constructed once (seed 20260811) from a mild factor structure and fixed for all datasets, conditions, and the true-performance test sample, so that each predictor had between-subject variance exactly ICC and total marginal variance exactly 1.0 (Cov_b = ICC × R), homogeneous across predictors. A binary outcome was generated for each cluster from a Bernoulli distribution with probability given by logit[P(Y_i = 1 | b_i)] = α + β^T b_i, using a sparse coefficient vector β (4 of 8 predictors nonzero, common log-odds magnitude |β| = 0.529, signs +, +, −, +, calibrated by pilot simulation to yield an oracle AUROC of 0.65 at the reference cell: ICC = 0.3, prevalence = 0.20, ρ = 0.5). Because the outcome depends only on the between-subject random effects b_i, whose distribution (Cov_b = ICC × R) does not involve ρ, the oracle AUROC is invariant to ρ by construction, so that ICC alone governs the true, between-subject signal available to the outcome model while ρ governs only the within-subject temporal structure of the observed predictor trajectories. The intercept was calibrated to the target prevalence at the population level by Gauss-Hermite quadrature (64 nodes), so that both the development sample and the independent test sample were draws from the same prespecified population. Each condition required exactly 500 valid datasets satisfying at least 3 events and 3 non-events; datasets failing this constraint were regenerated, with regeneration attempts, fitting errors, and fold-validity counts logged per condition.

#### Fitted model

The fitted design matrix comprised eight generated predictors together with a within-cluster day index (DOL), for nine columns in total. The day index carries no cluster-identifying information (it takes the same values 1 to 57 for every subject) and was included to match the motivating-dataset model structure; its presence is leakage- neutral with respect to subject identity.

#### True-performance reference

For each simulated dataset, the ridge model was refit on the full development data (all N subjects) and scored on a large independent test sample of new subjects (2,000 subjects) drawn from that dataset’s own population (identical factor covariance, coefficient vector, and calibrated intercept), generating 57- day AR(1) trajectories and aggregating to a subject-level prediction per test subject. The resulting AUROC estimated the true out-of-sample discrimination of the deployed model, providing the reference against which optimism was decomposed (Section 2.6).

#### Factorial design

Four factors were varied across a complete factorial design: cluster count N ∈ {20, 30, 50, 75, 100, 150}, intraclass correlation ICC ∈ {0.1, 0.3, 0.5}, lag-1 autocorrelation ρ ∈ {0.2, 0.5, 0.8}, and outcome prevalence ∈ {0.10, 0.20, 0.35}, yielding 162 combinations. Fixed parameters matched the motivating dataset: 57 observations per cluster and 8 generated predictors. The calibrated common coefficient magnitude (|β| = 0.529) was chosen to produce discrimination in the range observed in the motivating dataset (LOCO AUROC 0.608 to 0.650). For each combination, 500 datasets were generated, yielding 81,000 planned datasets (each condition required exactly 500 valid datasets; 81,000 total). Within each dataset, naive 10-fold, subject-level 10-fold, and subject-level 5-fold CV were each evaluated with 10 replicate fold assignments (outcome-stratified; all replicates required to be valid), LOCO was computed once, and the true-performance reference was computed once.

#### Signal-variance-controlled sensitivity arm

A signal-variance-controlled sensitivity analysis was conducted on a reduced grid of 24 conditions, comprising a core 18- condition grid (N ∈ {20, 30, 50, 75, 100, 150}, ICC ∈ {0.1, 0.3, 0.5}, ρ = 0.5, prevalence = 0.20) and a six-condition autocorrelation slice (N = 50, ICC ∈ {0.1, 0.3, 0.5}, ρ ∈ {0.2, 0.8}, prevalence = 0.20). Within this arm, coefficient magnitudes were scaled by √(ICC_ref / ICC), where ICC_ref = 0.3, holding the latent linear-predictor variance constant across ICC. This scaling holds the latent signal variance rather than the oracle AUROC itself constant, so a small gradient in true discrimination across ICC is expected to persist. Each of the 24 conditions was run under both a baseline and a controlled coefficient scale, with 500 datasets per condition per arm (24,000 dataset-arm rows in total). The two arms were paired by common random numbers (shared subject-effect, within-subject innovation, outcome, and fold-assignment draws), with only the coefficient scale and population-calibrated intercept differing between them; at ICC = 0.3 the scale factor equals one, so the arms coincide exactly there, providing a direct verification of the pairing. This design was intended to assess whether the apparent decline in the residual discrepancy with ICC reflects a genuine reduction in evaluation optimism or is partly attributable to the concurrent increase in oracle signal strength with ICC in the baseline arm, and whether the rise in the partitioning component with ICC is subject to the same confound. Because the sensitivity arm was executed as a separate run, its baseline arm constitutes an independent replication of the corresponding main-grid conditions rather than a reproduction of the identical draws.

Per-condition regeneration attempts, fitting errors, and fold-validity counts are reported in the reproducibility log (Appendix D). Of the 81,000 main-grid datasets, 14,703 required at least one regeneration (18.2%), concentrated in conditions with N = 20 and 10% prevalence; zero datasets were flagged and zero fit errors were recorded across all 162 conditions. Ridge logistic regression with λ = 0.01 and balanced class weights was used throughout, matching the motivating dataset.

### 2.8 Use of generative artificial intelligence tools

Claude Opus 4.7 (Anthropic, San Francisco, CA, USA) was used to assist with refinement of selected R code, troubleshooting, and language editing of portions of the manuscript. The author designed the study, specified the simulation framework, conducted and verified all analyses, drafted the manuscript with AI writing assistance, reviewed and edited all AI-generated content, and takes full responsibility for the integrity, accuracy, and interpretation of the work. No generative artificial intelligence tool was listed as an author, used to generate original research data, or used to make autonomous analytic decisions.

## 3. Results

### 3.1 Motivating dataset: observation-level CV comparison

Table 1 presents the CV comparison for severe ROP. Naive observation-level 10-fold CV yielded a mean subject-level AUROC of 0.686 across 50 replicate fold assignments, LOCO produced an AUROC of 0.608 (subject-level bootstrap 95% CI: 0.461, 0.745), and subject-level 10-fold CV produced a mean AUROC of 0.592. The optimism of naive CV relative to LOCO was +0.078 AUROC units, and subject-level 10-fold CV deviated from LOCO by −0.015. The interpretive consequence is notable: naive CV conveys a substantially more favorable impression of discrimination, whereas the subject-level bootstrap interval for the LOCO AUROC remains compatible with chance performance. The subject-level bootstrap interval for the LOCO AUROC nearly coincided with the DeLong interval (0.464, 0.752), so the choice of interval method does not alter any substantive conclusion. The bootstrap is reported because it does not assume that all predictions arise from a single fixed model.

**Table 1.**
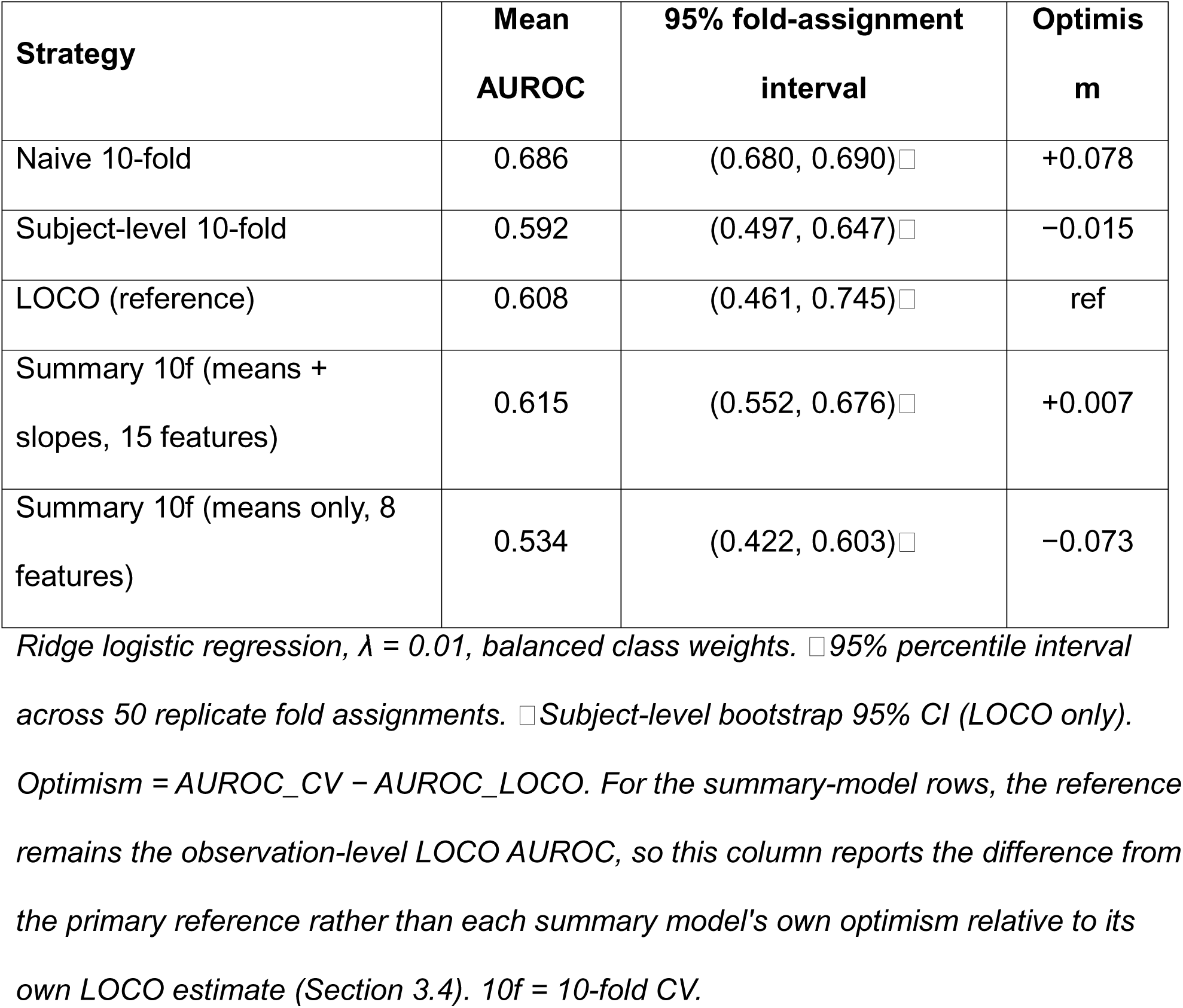
Cross-validation comparison for severe ROP prediction (N = 101 subjects, 15 events, 5615 daily observations).

Table 2 presents any ROP. The optimism of naive CV was +0.031 (0.682 versus 0.650 for LOCO; subject-level bootstrap 95% CI: 0.545, 0.753), and subject-level 10-fold CV deviated from LOCO by −0.000. The smaller optimism for any ROP is consistent with its larger effective sample size (48 versus 15 events), and all strategies exceeded chance discrimination.

**Table 2.** Cross-validation comparison for any ROP prediction (N = 101 subjects, 48 events, 5615 daily observations).

| Strategy | Mean AUROC | 95% fold-assignment interval | Optimism |
| --- | --- | --- | --- |
| Naive 10-fold | 0.682 | (0.680, 0.683)□ | +0.031 |
| Subject-level 10-fold | 0.650 | (0.626, 0.669)□ | −0.000 |
| LOCO (reference) | 0.650 | (0.545, 0.753)□ | ref |
| Summary 10f (means + slopes, 15 features) | 0.579 | (0.490, 0.633)□ | −0.071 |
| Summary 10f (means only, 8 features) | 0.620 | (0.575, 0.661)□ | −0.030 |

*Footnotes as in Table 1.*

### 3.2 Motivating dataset: coefficient stability

To characterize estimation stability at low events-per-variable (EPV), the standardized ridge coefficients were recorded across all subject-level training folds (10 folds × 10 replicate assignments = 100 folds; Table 3). For severe ROP (EPV ≈ 1.7), the dominant predictors were sign-stable across folds with moderate variability (titration index, mean 0.299, SD 0.084; Avg_FiO , 0.221, SD 0.088; ambient hyperoxemia, −0.190, SD 0.079; Swings, 0.154, SD 0.067; iatrogenic hyperoxemia, −0.145, SD 0.049), whereas near-null predictors changed sign across folds (severe hypoxemia, 0.027, SD 0.055). For any ROP (EPV ≈ 5.3), coefficients were markedly more stable (DOL, 0.177, SD 0.018; Avg_FiO , 0.305, SD 0.053; ambient hyperoxemia, −0.308, SD 0.051). The improvement in stability with additional events is the expected consequence of low EPV and indicates that individual coefficient magnitudes in the severe ROP model should be interpreted with caution, while the discriminative comparison of CV strategies is unaffected. The two outcomes also differed in which predictors dominated the model, most notably Swings (severe ROP mean 0.154, any ROP mean −0.002), so the improved stability at higher EPV should not be read as recovering a consistent set of dominant predictors across outcomes.

**Table 3.**
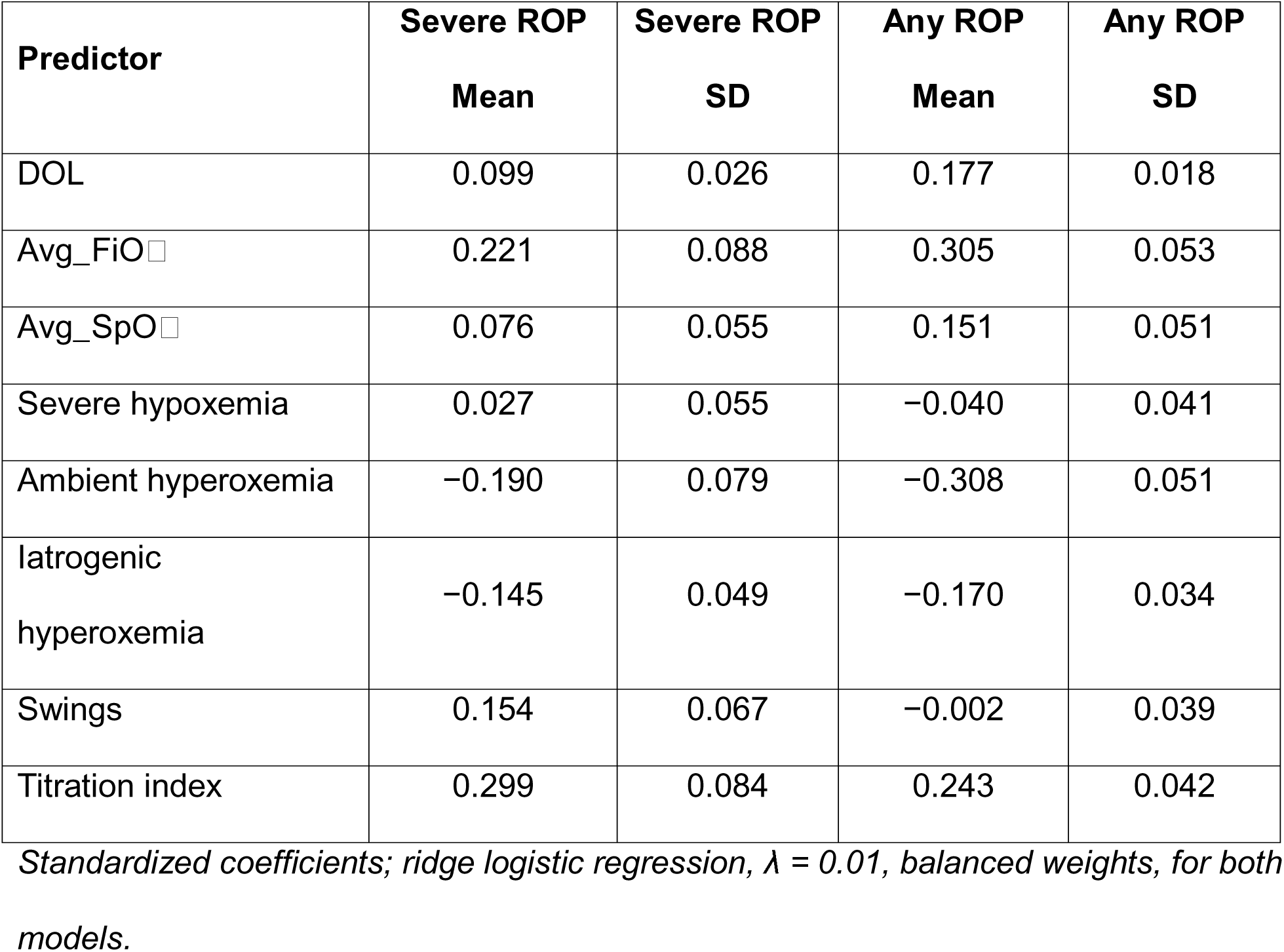
Fold-wise standardized coefficient stability by ROP severity (subject-level 10-fold CV, 100 folds).

### 3.3 Motivating dataset: precision illusion

Naive CV produced not only biased but artificially precise estimates. For severe ROP, the standard deviation of naive 10-fold AUROC across replicate fold assignments was 0.0026, compared with 0.0351 for subject-level 10-fold CV, an approximately 13-fold difference; for any ROP the values were 0.0011 and 0.0110, an approximately 10-fold difference. This precision illusion arises because the approximately 5,600 non-independent daily observations are treated as if they were 5,600 independent units, markedly inflating the effective sample size.

### 3.4 Motivating dataset: subject-level summary comparison

Aggregating daily observations to per-subject summaries removed the source of within-observation dependence but at the cost of temporal information, and the result depended on outcome and feature count. For severe ROP, the means-plus-slopes summary (15 features) achieved a LOCO AUROC of 0.619, slightly exceeding the observation-level LOCO of 0.608, whereas the means-only summary (8 features) achieved only 0.434, well below; the marked gap between the means-only 10-fold estimate (0.534) and its LOCO counterpart (0.434, subject-level bootstrap 95% CI 0.402 to 0.719) reflects LOCO instability at extremely low EPV (≈ 1.9) rather than systematic bias, as leaving out a single event subject meaningfully alters the fitted model. For any ROP, all summary configurations performed at or below the observation-level LOCO (0.650), consistent with overfitting at low EPV. These results indicate that subject-level aggregation can mitigate leakage but discards within-subject temporal structure, whereas observation-level modeling with cluster-aware CV retains the observation-level information while avoiding leakage.

### 3.5 Motivating dataset: regularization sensitivity

The optimism of naive CV relative to LOCO persisted across a 100,000-fold range of regularization strength. For severe ROP, the optimism was +0.051 at strong regularization (λ = 1.0), +0.078 at moderate (λ = 0.01), and +0.096 at weak (λ = 0.00001); for any ROP, the corresponding values were +0.025, +0.031, and +0.032. The optimism therefore arises fundamentally from the partitioning scheme rather than model complexity, although its magnitude increased modestly with weaker regularization.

### 3.6 Simulation study: the partitioning component by cluster count

Table 4 presents the partitioning component (naive minus subject-level CV) aggregated by cluster count. It was substantial at every sample size, decreasing monotonically from +0.204 at N = 20 to +0.104 at N = 150. Even at the largest sample size the partitioning optimism exceeded 10 AUROC points, large on the AUROC scale.

**Table 4.**
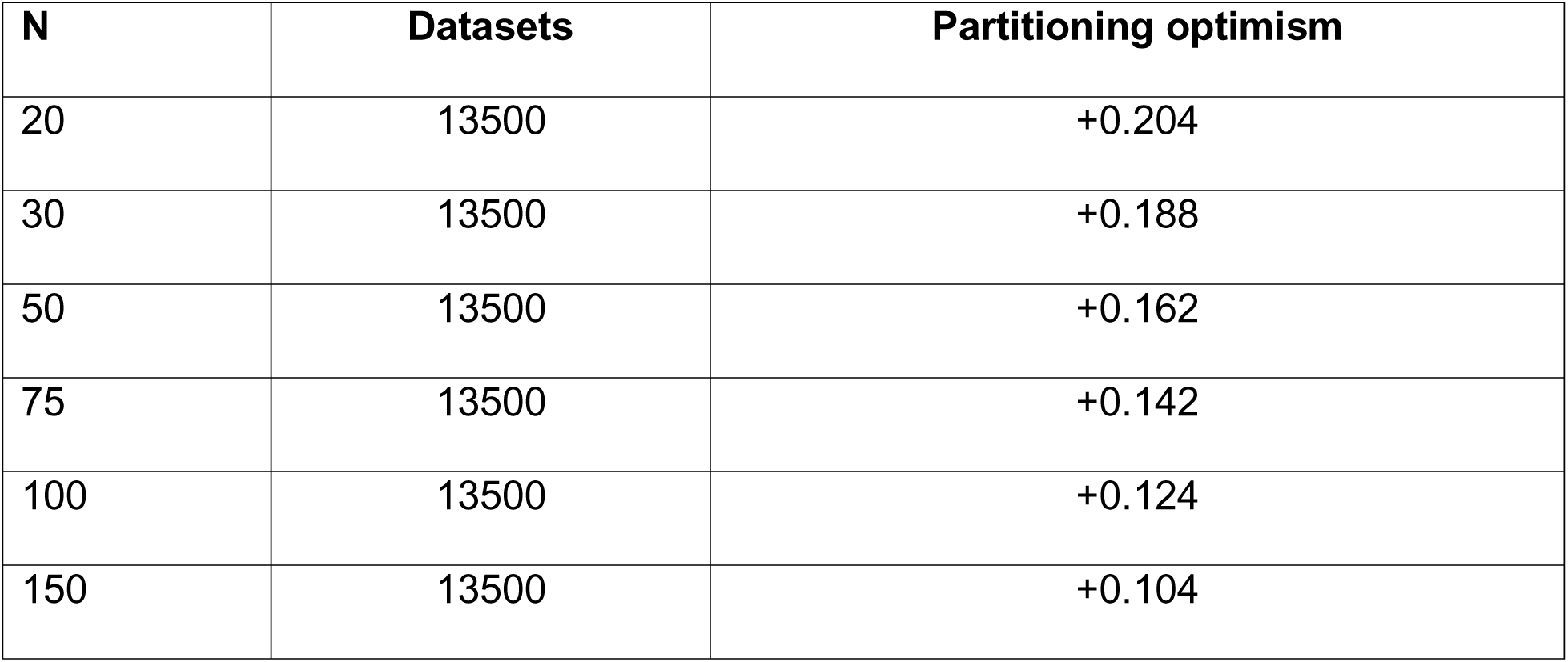
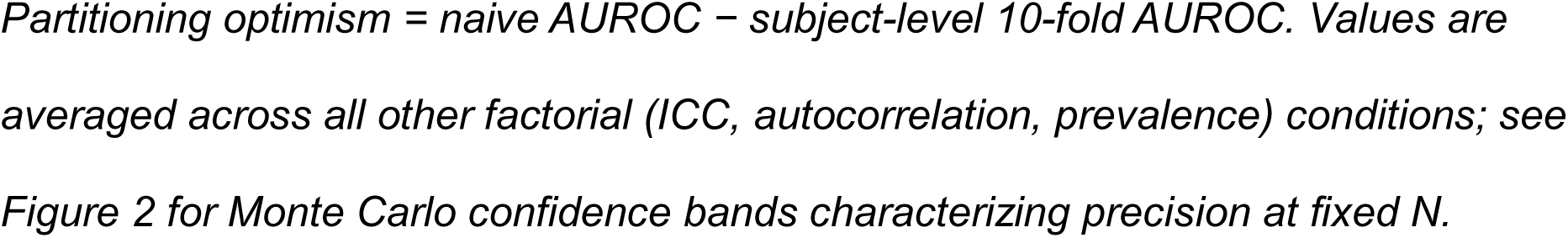
Partitioning component of optimism by cluster count, averaged across ICC, autocorrelation, and prevalence.

| N | Datasets | Partitioning optimism |
| --- | --- | --- |
| 20 | 13500 | +0.204 |
| 30 | 13500 | +0.188 |
| 50 | 13500 | +0.162 |
| 75 | 13500 | +0.142 |
| 100 | 13500 | +0.124 |
| 150 | 13500 | +0.104 |

### 3.7 Simulation study: effects of ICC, autocorrelation, and prevalence

Each remaining factor influenced the partitioning component in the expected direction. Higher ICC increased it (mean +0.125 at ICC = 0.1, +0.161 at 0.3, +0.175 at 0.5); higher autocorrelation increased it (+0.150 at ρ = 0.2, +0.152 at 0.5, +0.160 at 0.8); and lower prevalence increased it (+0.176 at 10%, +0.151 at 20%, +0.134 at 35%). Figure 2 illustrates the joint dependence on cluster count and ICC across autocorrelation and prevalence.

**Figure 2.**
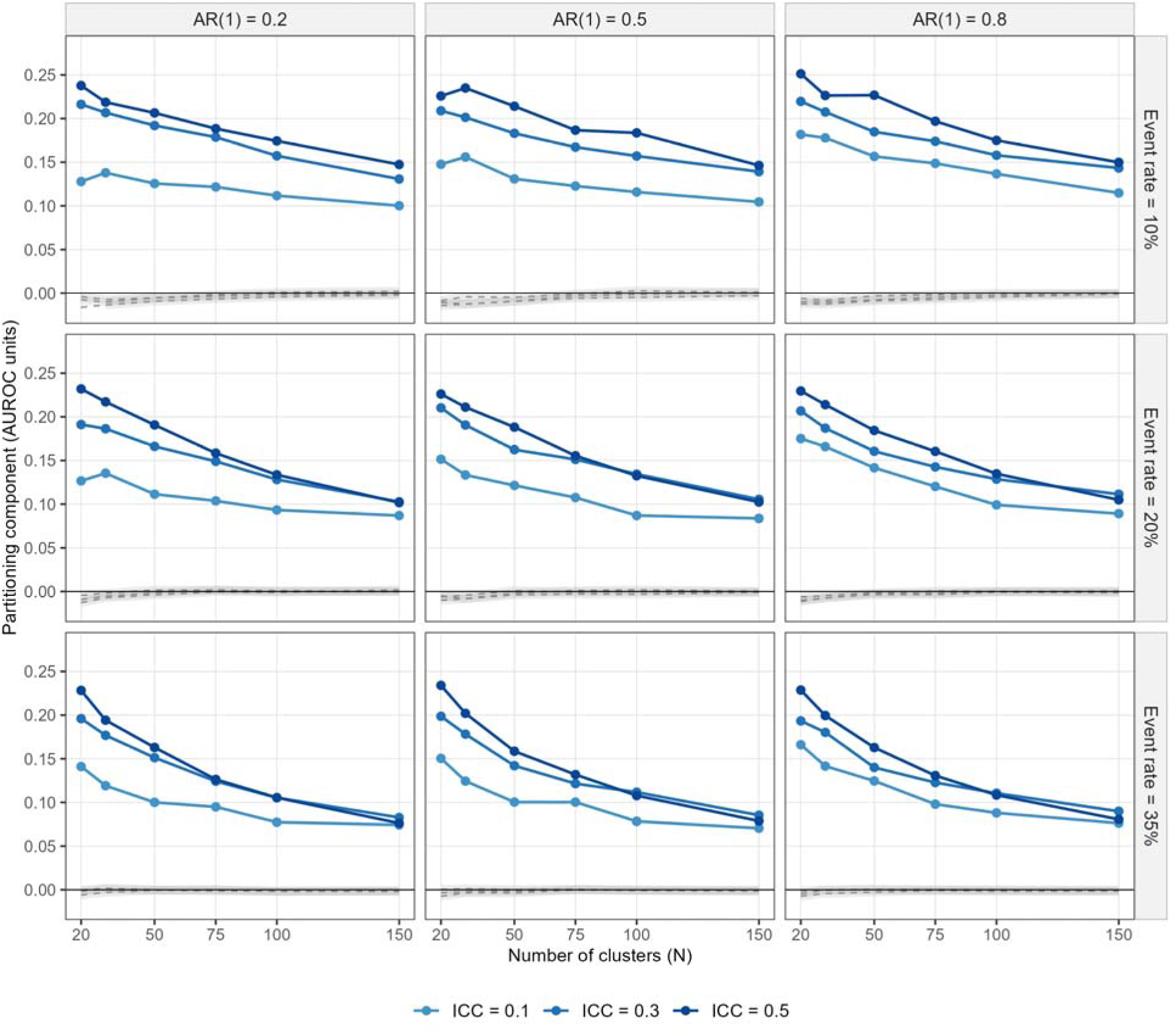
Partitioning component of optimism by cluster count and ICC. Solid lines show the partitioning component by number of clusters (N) and ICC, paneled by lag-1 autocorrelation (columns) and outcome prevalence (rows). Dashed grey lines show subject-level 10-fold CV deviation from LOCO (with 95% Monte Carlo confidence bands), near zero across all 162 conditions. Quantities are in AUROC units.

Within the partitioning component, an informative interaction was observed between ICC and autocorrelation (Table 5). At low ICC (0.1), increasing autocorrelation from 0.2 to 0.8 increased the partitioning optimism modestly, by 0.023, smaller than the ICC main effect’s own 0.050 range across this factor; the effect was smaller still at ICC = 0.3 and 0.5 (Table 5), consistent with the 21% relative increase at ICC = 0.1 versus 3% at ICC = 0.5 reported in Appendix C. Qualitatively, when between-cluster variation is already high, clusters are readily distinguishable by their stable predictor levels and the marginal contribution of temporal autocorrelation is small. When between-cluster variation is low, temporal autocorrelation becomes the primary route by which naive partitioning exploits within-subject similarity. This interaction concerns the internal structure of the partitioning component and is distinct from the decomposition of total optimism presented in Section 3.9. A related N-by-ICC attenuation is visible in Figure 4: the sensitivity of the partitioning component to ICC is most pronounced at small cluster counts and diminishes progressively as N increases, consistent with the interpretation that larger samples allow naive partitioning to exploit between-cluster signal through stable predictor levels regardless of ICC level.

**Figure 3.**
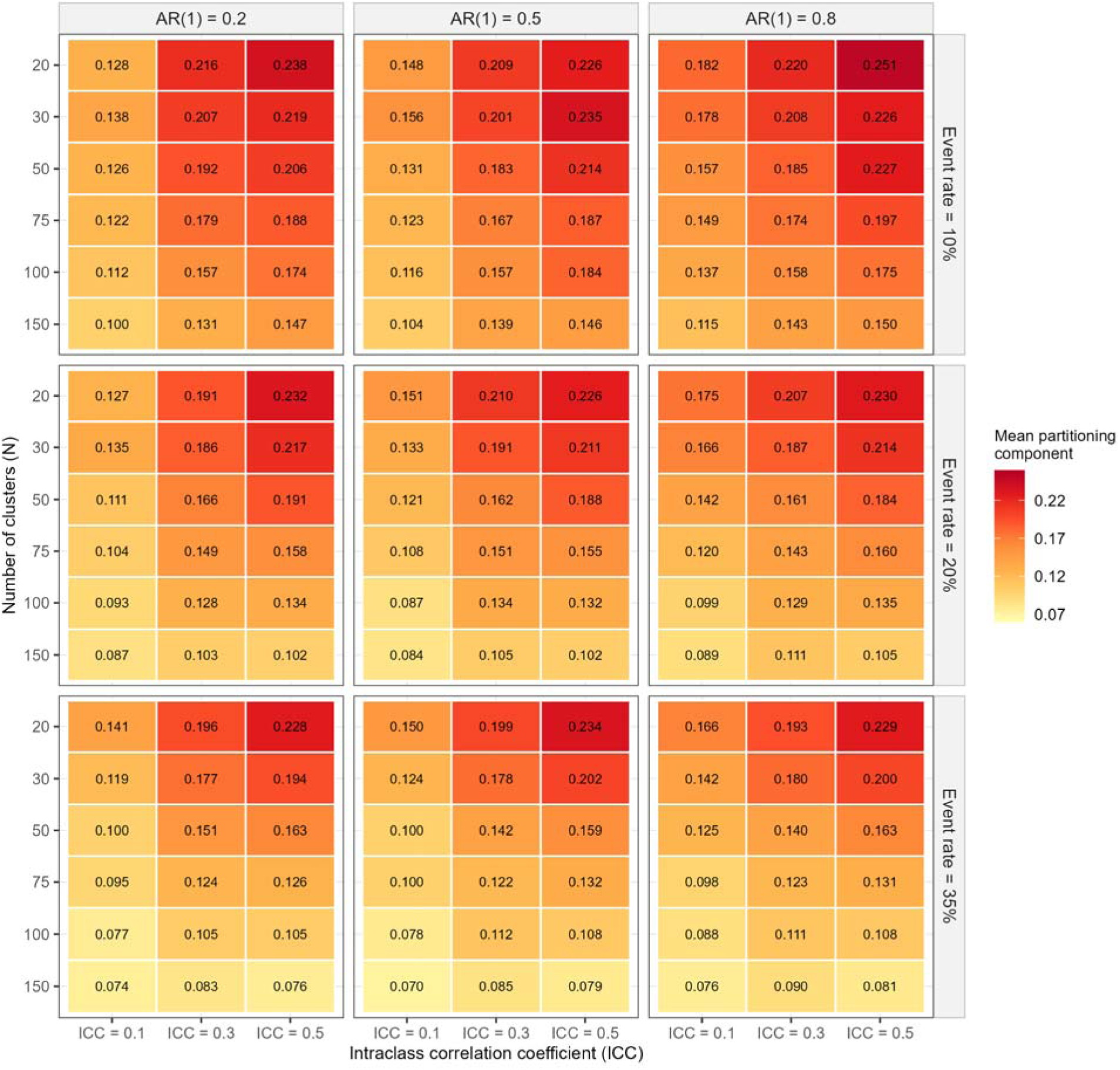
Heat map of the partitioning component of optimism (AUROC units) across all 162 conditions. Rows represent cluster count, columns represent ICC, with panels organized by lag-1 autocorrelation (columns) and prevalence (rows).

**Figure 4.**
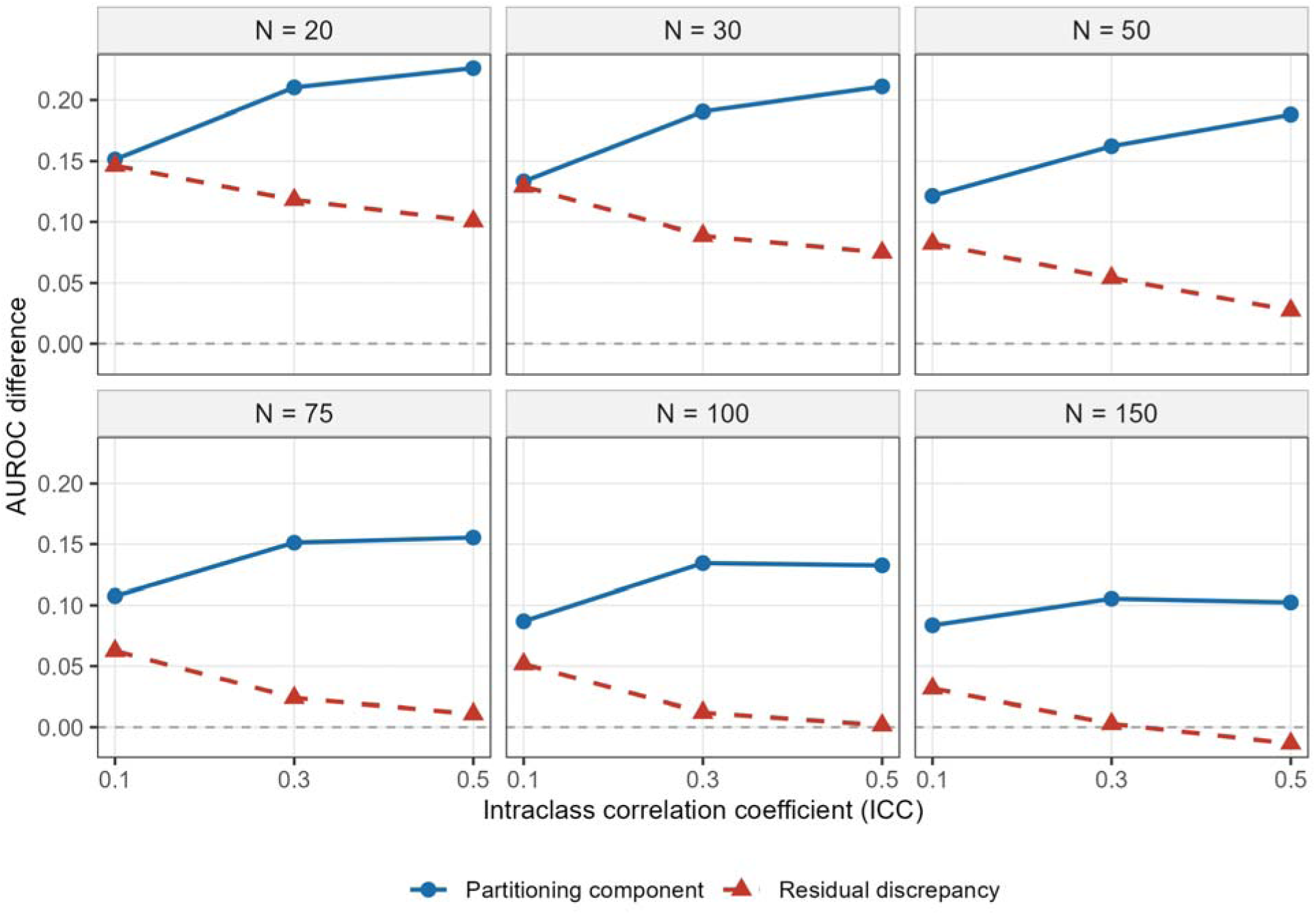
The dependence-axis dissociation. Within each cluster-count stratum, the partitioning component of optimism (naive minus subject-level CV) rises with ICC while the residual discrepancy (subject-level CV minus true performance) falls with ICC, the two moving in opposite directions on the dependence axis. Quantities are in AUROC units.

**Table 5.** Partitioning component by ICC and lag-1 autocorrelation (ρ).

| ICC | $\rho = 0.2$ | $\rho = 0.5$ | $\rho = 0.8$ |
| --- | --- | --- | --- |
| 0.1 | +0.115 | +0.121 | +0.139 |
| 0.3 | +0.161 | +0.161 | +0.162 |
| 0.5 | +0.173 | +0.175 | +0.178 |

### 3.8 Simulation study: subject-level CV performance

Subject-level 10-fold CV closely agreed with LOCO across all 162 conditions (condition-level mean absolute deviation 0.004, maximum 0.016), maintained at sample sizes as small as N = 20, where only two subjects are held out per fold. Subject-level 5-fold CV showed similarly close agreement (mean absolute deviation 0.002, maximum 0.009) with greater replicate variability, as expected with larger held-out fractions. The precision illusion documented in the motivating dataset was confirmed across all conditions: the ratio of naive to subject-level 10-fold CV standard deviation had a median of 0.095 (range 0.045 to 0.358).

### 3.9 Simulation study: true-performance reference and the dependence-axis dissociation

Referencing performance against a large independent sample of new subjects located naive CV, subject-level CV, and LOCO relative to true out-of-sample discrimination.

Averaged over all conditions, naive CV AUROC was 0.779, subject-level 10-fold and LOCO were 0.625 and 0.628, and true AUROC was 0.566. Subject-level CV and LOCO thus coincided with each other and tracked true performance to within the residual discrepancy, while naive CV lay well above both. The total optimism of naive CV relative to truth (mean +0.213, range +0.068 to +0.363) was split into a partitioning component (mean +0.154, range +0.071 to +0.257) and a residual discrepancy (mean +0.059, range −0.013 to +0.170), the latter shared by subject-level CV and LOCO alike. Small negative values of the residual discrepancy at the largest cluster counts reflect conditions in which the full-data model and the cross-validated model are nearly equivalent, and sampling variance in the test sample produces estimates near zero.

This split is definitional once a true-performance reference is available, since the two components are constructed to sum to total optimism; its scientific content lies not in the arithmetic but in the distinct empirical behavior of the two resulting quantities, examined next.

The two components dissociated along the dependence axis. The partitioning component rose with within-subject dependence, increasing from +0.115 at the lowest dependence corner (ICC = 0.1, ρ = 0.2; N = 50, 20% prevalence) to +0.186 at the highest dependence corner (ICC = 0.5, ρ = 0.8), a 61% increase. Because oracle AUROC also rises with ICC in this arm (from 0.528 to 0.603 on average), part of this contrast reflects signal strength rather than dependence; the signal-variance-controlled arm separates the two directly (Section 3.10). The residual discrepancy did not rise with dependence: it was approximately flat across autocorrelation (+0.055, +0.058, +0.063 across ρ = 0.2, 0.5, 0.8) and declined with ICC (+0.086, +0.055, +0.036 across ICC = 0.1, 0.3, 0.5), moving in the opposite direction to the partitioning component. The proposed explanation for the ICC decline is that higher ICC raises the true between-subject signal and thereby leaves less room for evaluation optimism, an explanation the sensitivity arm tests directly. The opposite response to ICC held within every cluster-count stratum without exception (for example, at N = 50 with ρ = 0.5 and 20% prevalence, the partitioning component rose +0.125 → +0.164 → +0.187 across ICC = 0.1, 0.3, 0.5, while the residual discrepancy fell +0.083 → +0.054 → +0.027). Both components declined with cluster count and with prevalence, as any optimism should, so that the two are positively correlated across conditions overall; the dissociation is specifically on the dependence axis. That the partitioning component rises steeply and monotonically with ICC while the residual discrepancy does not rise at all establishes the partitioning optimism as a distinct, dependence-driven mechanism rather than a relabeling of the residual discrepancy that the cluster-aware CV schemes examined always carry. Figure 4 displays this dissociation.

To confirm that the residual discrepancy is not an artifact of the particular fold ratio used, the true-performance comparison was repeated for subject-level 5-fold CV, which trains on a smaller fraction of subjects per fold (80%) than 10-fold (90%) or LOCO (nearly the full sample). The residual discrepancy at K = 5 was materially indistinguishable from that at K = 10: mean +0.057 across conditions, versus mean +0.059 at K = 10, and mean +0.062 for LOCO. The K = 5 response to dependence reproduced the K = 10 pattern as well, declining with ICC (+0.084, +0.053, +0.034 across ICC = 0.1, 0.3, 0.5, versus +0.086, +0.055, +0.036 at K = 10) and remaining approximately flat across autocorrelation (+0.053, +0.056, +0.061 across ρ = 0.2, 0.5, 0.8, versus +0.055, +0.058, +0.063 at K = 10). The condition-level correlation between the K = 5 and K = 10 residual discrepancies was 0.9994. Because reducing the per-fold training fraction from 90% to 80% produced no detectable change in the residual discrepancy, it appears governed by the size of the development sample as a whole rather than by the training-to-test ratio within each fold, a distinction not resolvable from the K = 10-versus-LOCO comparison alone.

### 3.10 Simulation study: signal-variance-controlled sensitivity arm

The 24-condition sensitivity arm isolated the contribution of the ICC-to-signal confound. The paired arms coincided exactly at ICC = 0.3, where the coefficient scale factor equals one, confirming the common-random-number pairing. Signal control reduced the ICC gradient in true discrimination by 78%, from 0.076 to 0.017 AUROC units across ICC = 0.1 to 0.5, but did not eliminate it, as anticipated in Section 2.7. Condition- weighted aggregates were essentially unchanged between arms (total optimism +0.209 baseline versus +0.210 controlled; partitioning component +0.155 versus +0.158; residual discrepancy +0.054 versus +0.052), as were scheme invariance (mean |LOCO − subject-level 10-fold| = 0.003 in both arms) and the precision illusion (median standard deviation ratio 0.103 in both arms).

The two components responded differently to the control (Table 6). The rise in the partitioning component across ICC was largely retained, from +0.052 in the baseline arm to +0.041 under control, and remained monotone within all six cluster-count strata. At the dependence corners (N = 50), it rose from +0.122 to +0.197 in the baseline arm and from +0.145 to +0.192 under control. The decline in the residual discrepancy across ICC was attenuated by 73%, from −0.049 to −0.013, and remained monotone in only four of six strata. Its relationship to development-sample size was unaffected (+0.116, +0.090, +0.054, +0.027, +0.015, +0.006 across N = 20 to 150 under control, versus +0.114, +0.091, +0.056, +0.029, +0.018, +0.012 in the baseline arm; correlation with log N = −0.97 under control). The baseline arm of this run reproduced the corresponding main-grid conditions closely (for example, partitioning component +0.124 versus +0.125 and residual discrepancy +0.087 versus +0.081 at N = 50, ICC = 0.1, ρ = 0.5), providing an independent replication of the main grid.

**Table 6.** Baseline and signal-variance-controlled arms by ICC.

| ICC | Partitioning,<br>baseline | Partitioning,<br>controlled | Residual,<br>baseline | Residual,<br>controlled | True AUROC,<br>baseline | True AUROC,<br>controlled |
| --- | --- | --- | --- | --- | --- | --- |
| 0.1 | +0.122 | +0.131 | +0.080 | +0.059 | 0.529 | 0.557 |
| 0.3 | +0.160 | +0.160 | +0.048 | +0.048 | 0.570 | 0.570 |
| 0.5 | +0.174 | +0.172 | +0.032 | +0.046 | 0.605 | 0.575 |

*Core 18-condition grid ( $\rho = 0.5$ , prevalence = 0.20), condition-weighted equal means across the six cluster counts; 500 datasets per condition per arm. Arms are paired by common random numbers and coincide exactly at $ICC = 0.3$ , where the coefficient scale factor equals one. Partitioning = naive AUROC – subject-level 10-fold AUROC; residual = subject-level 10-fold AUROC – true AUROC.*

## 4. Discussion

This study quantified and decomposed the optimism produced by naive observation-level cross-validation in a clinically common but methodologically underexamined data structure: continuous repeated-measures predictors paired with a binary subject-level outcome. Naive CV produced substantial, systematic overestimates of discriminative performance across a broad range of realistic conditions, and simple subject-level partitioning eliminated the leakage-driven component of that optimism, yielding estimates that agreed closely with leave-one-cluster-out validation and whose remaining discrepancy from true out-of-sample performance decreased strongly with cluster count.

### 4.1 Two components of optimism

The central result is that the total optimism of naive CV in this setting is the sum, by construction, of two components that prove to have mechanistically distinct determinants. The partitioning component, the difference between naive and cluster-aware CV, is caused by within-subject leakage and is what subject-level partitioning removes; it ranged from +0.071 to +0.257 and rose with within-subject dependence. The residual discrepancy, the difference between cluster-aware CV and the true AUROC of the full-data model, is governed primarily by development-sample size and not by within-subject dependence; it declined with cluster count and did not increase with dependence. Because naive and subject-level CV share the identical residual discrepancy, they differ only in the partitioning component, and cluster-aware partitioning removes that component in full. None of the cluster-aware CV schemes examined removes the residual discrepancy; reducing it requires more data or explicit optimism correction, such as the optimism-corrected bootstrap [20,21] applied at the cluster level. The dependence-axis dissociation, in which the partitioning component rises steeply with ICC while the residual discrepancy does not rise at all, provides direct evidence that the partitioning optimism is a genuine, dependence-driven mechanism and not a restatement of the residual discrepancy that the cluster-aware CV schemes examined always carry. The paired signal-variance-controlled arm confirmed that the partitioning component’s response to ICC is not an artifact of the concurrent rise in oracle discrimination, whereas most of the residual discrepancy’s apparent decline with ICC is attributable to that rise, so that the residual is more purely governed by development-sample size than the baseline arm alone indicated (Section 3.10).

The worst-case simulation condition (N = 20, ICC = 0.5, ρ = 0.8, prevalence = 0.10) produced a mean partitioning component reaching +0.257, large enough in principle to materially change the apparent clinical utility of a model: in that condition, the true AUROC was 0.569, indicating discrimination only modestly better than chance, whereas naive CV returned a mean AUROC of 0.932, appearing highly discriminative, and even the most favorable condition (N = 150, ICC = 0.1, ρ = 0.5, prevalence = 0.35) retained a partitioning component of +0.071. In the motivating dataset, the +0.078 optimism for severe ROP represented the difference between a model that appears somewhat useful (AUROC 0.686) and one with limited discrimination (0.608), and it reversed the qualitative conclusion about whether the model discriminates better than chance.

### 4.2 The ICC by autocorrelation interaction

Within the partitioning component, the interaction between ICC and autocorrelation is informative about how naive partitioning exploits within-subject similarity. At low ICC, temporal autocorrelation is the dominant route to leakage, because (1 − ICC) is exactly the within-subject variance share on which ρ can act (Appendix C). At high ICC, clusters are already distinguishable by their stable predictor levels and the marginal effect of autocorrelation is small, since little within-subject variance remains for ρ to structure.

This pattern indicates that ICC and autocorrelation, although orthogonal as data-generating parameters, combine in a partially redundant rather than additive fashion: once between-cluster heterogeneity is sufficient to make clusters identifiable from any single observation, additional temporal structure adds comparatively little further leakage. Prediction models built on data with high between-cluster heterogeneity are therefore vulnerable to naive CV optimism regardless of temporal structure, whereas models built on more homogeneous populations require substantial temporal autocorrelation for large optimism. This structure is presented qualitatively; a formal separation of the two dependence pathways, including the limiting case of time-invariant covariates, is the subject of ongoing follow-up work.

### 4.3 Subject-level CV as a practical remedy

Subject-level 10-fold CV closely agreed with LOCO across all conditions, including at N = 20, which is encouraging for the many clinical settings with small cluster counts, such as neonatal intensive care, rare-disease registries, and smaller single-institution studies. LOCO is deterministic and therefore has no fold-assignment distribution, so characterizing its uncertainty requires a separate inferential procedure such as the subject-level bootstrap used here, whereas subject-level k-fold CV yields a distribution of AUROC estimates across repeated random partitions that characterizes partitioning variability directly. Inference for cross-validated AUROC under within-subject clustering was not addressed in this study. Subject-level k-fold CV is therefore the recommended default, with LOCO reserved as a bias reference. This recommendation requires no new methodology. Subject-level partitioning is implemented in standard software (for example, the group_vfold_cv function in the rsample R package [22]).

A corollary concerns hyperparameter tuning. When regularization or other hyperparameters are selected by nested CV, the partitioning must be performed at the subject level at every level of nesting. Naive inner folds leak during tuning exactly as naive outer folds leak during evaluation, so that naive nested tuning both inflates the performance estimate and can mis-select the hyperparameter. Cluster-aware partitioning must therefore be applied to inner and outer folds alike.

### 4.4 Relationship to true performance

By referencing performance against a large sample of new subjects, this study distinguished what cluster-aware partitioning does and does not accomplish. It removes the leakage-driven optimism entirely, but the residual discrepancy that the cluster-aware CV schemes examined here carry remains, and its magnitude in this setting was appreciable at small cluster counts (averaging +0.119 at N = 20). Whether cluster-level optimism-corrected bootstrap validation reduces this residual discrepancy was not evaluated here. Any such procedure should resample subjects with replacement rather than observations [20,21], since observation-level resampling reproduces the leakage that cluster-aware partitioning is intended to remove. Because the cross-validated models were trained on fewer subjects than the full-data model against which the residual is defined, and because true discrimination improved with development-sample size on average across the conditions examined (mean true AUROC rising from 0.551 at N = 20 to 0.584 at N = 150), the training-size mismatch biases this residual downward, so the positive values observed cannot be attributed to that mismatch. The residual discrepancy’s relationship to development-sample size was also unaffected by the internal fold ratio (Section 3.9) and by signal-variance control (Section 3.10), indicating that this relationship reflects the size of the development sample rather than how that sample is partitioned or the strength of the underlying signal, and that neither LOCO nor a choice among reasonable values of k for subject-level k-fold CV offers a path to reducing the residual on that basis. The partitioning principle, cluster-level rather than observation-level, applies to all resampling-based validation strategies.

### 4.5 Subject-level summary approaches

The comparison with subject-level summaries yielded a nuanced result: for severe ROP, the means-plus-slopes summary slightly outperformed the observation-level model on LOCO AUROC (0.619 versus 0.608), while for any ROP the observation-level model with cluster-aware CV outperformed all summary configurations. Aggregation avoids leakage by removing dependence, whereas cluster-aware CV retains the observation-level information while avoiding leakage. Subject-level feature engineering should therefore be viewed as a complement to, rather than a replacement for, cluster-aware partitioning.

### 4.6 Prevalence of naive CV in clinical prediction studies

The prevalence of observation-level CV on longitudinal data warrants emphasis. Previously reported AUROC estimates from NICU vital-sign prediction studies using standard k-fold CV without documented subject-wise partitioning [23–25] may be optimistically biased to a degree that depends on the cluster count, ICC, autocorrelation, and prevalence of each cohort; notably, one modeling-competition entry that did employ subject-aware cross-validation achieved the best-generalizing performance among five otherwise-comparable approaches [23]. That the choice of CV technique alone can materially alter reported performance in machine-learning-based diagnostic applications has been demonstrated previously [26]. The present results quantify how large that alteration becomes in this specific data structure, reaching +0.257 AUROC units at the upper end of the simulated space, a magnitude sufficient in principle to change the interpretation of a model from clinically useful to non-discriminating.

### 4.7 Scope and generalizability

The two leakage pathways characterized here are properties of the partitioning scheme relative to the data structure, not of the binary outcome or the AUROC metric specifically. The partitioning principle, that naive observation-level CV is optimistically biased whenever repeated observations from the same subject can appear in both training and test partitions, is therefore expected to extend to continuous, count, and time-to-event outcomes, although the magnitude of the resulting optimism and its behavior on other performance metrics would require separate evaluation. A recent broad overview reached this general conclusion for clustered data [11]. The present study supplies what that general literature does not, namely the first factorial quantification of the magnitude as a function of clinically measurable parameters for the single structure, continuous repeated-measures predictors paired with a subject-level binary outcome evaluated by AUROC, that is especially common in settings employing continuous physiological monitoring, such as neonatal care, critical care, and wearable-sensor research. This is a deliberate scope decision. A study spanning every outcome type and metric would sacrifice the parameter-level resolution that is this paper’s principal contribution.

Further methodological extensions to this data structure, including alternative leakage pathways, calibration, and sample-size planning, are left to future work.

### 4.8 Limitations

Several limitations should be noted. First, the simulation employed a homogeneous dependence structure, applying the same ICC and autocorrelation to all predictors, whereas the motivating dataset showed predictor ICCs from 0.16 to 0.53 and lag-1 autocorrelations from 0.45 to 0.73. Heterogeneous predictor dependence is an identified priority for future work. Second, the factorial design included no zero- dependence cell (the lowest dependence examined was ICC = 0.1, ρ = 0.2), so the partitioning component is shown to decrease monotonically toward the low-dependence corner rather than to vanish at exactly zero dependence. The pure-autocorrelation limit is treated in ongoing follow-up work, together with the fixed-covariate limiting case (ICC = 1). Third, the motivating analysis used a single dataset from a single institution, and replication in other domains would strengthen external validity. Fourth, the prediction model was ridge logistic regression. Although the regularization sensitivity analysis showed that optimism persists across a wide range of λ, models with greater capacity to memorize within-cluster structure, such as gradient-boosted trees or neural networks, may exhibit larger partitioning optimism, so the magnitudes reported here are plausibly conservative for more flexible learners. This restriction to a single, well-characterized learner is a deliberate scope decision, made to preserve the parameter-level resolution across ICC, autocorrelation, cluster count, and prevalence that constitutes the present study’s principal contribution. Learner-dependence in this and related leakage mechanisms is the subject of ongoing follow-up work. Fifth, AUROC was the sole metric. Whether naive CV bias extends comparably to calibration measures is left to future work. Sixth, the signal-variance control equalized the latent linear-predictor variance rather than the oracle AUROC itself, so a modest gradient in true discrimination across ICC (0.557 to 0.575) persisted within the controlled arm, and that arm covered 24 of the 162 conditions at a single prevalence, so its conclusions are best read as applying to the ICC contrast it was designed to isolate. Seventh, this study did not examine external validation in an independent dataset, which remains the most rigorous test of generalizability [5,16]. Eighth, the requirement that each simulated dataset contain at least three events and three non-events, imposed to permit stable model fitting, inflated the realized subject-level prevalence above the nominal target in the smallest samples at the lowest prevalence, the cells in which regeneration was concentrated (Section 2.7). The mean realized prevalence at N = 20 was 0.18 at a nominal 0.10 and 0.23 at a nominal 0.20, and at N = 30 was 0.14 at a nominal 0.10, whereas the realized and nominal rates coincided to within 0.01 for all conditions with N ≥ 50 (Appendix D). The primary naive versus cluster-aware comparison is unaffected by this conditioning, as both estimates were computed on the same accepted datasets. Because optimism was found to increase as prevalence decreased, the conditioning attenuated rather than exaggerated the optimism reported in the affected small-sample cells, so the monotonic decline of the partitioning component with increasing sample size would be expected to steepen, not reverse, in its absence.

### 4.9 Summary of contributions

To the author’s knowledge, this study provides the first factorial quantification of naive CV optimism on the AUROC scale, referenced against true out-of-sample performance, for the repeated-measures predictor, subject-level binary outcome data structure, mapping its magnitude across 162 clinically relevant conditions and decomposing it into a dependence-driven partitioning component and a sample-size-governed residual internal-validation discrepancy with dissociable signatures on the dependence axis. A paired signal-variance-controlled arm establishes that the dependence-driven behavior of the partitioning component is not an artifact of the covariation between ICC and true discrimination. The study documents a precision illusion in which naive CV produces artificially narrow AUROC distributions, and it confirms subject-level 10-fold CV as a practical substitute for LOCO, in close agreement across all conditions examined, including sample sizes as small as N = 20.

## 5. Conclusions

Naive observation-level CV produces systematic and meaningful overestimates of discriminative performance when continuous predictors are measured repeatedly within subjects and the binary outcome is defined at the subject level. The leakage-driven partitioning component of this optimism ranged from +0.071 to +0.257 across the parameter space examined, was larger under greater within-subject dependence, and was accompanied by a precision illusion that made the biased estimates appear artificially stable. Subject-level CV partitioning eliminates this component and agrees closely with leave-one-cluster-out validation, leaving a residual internal-validation discrepancy that was observed across all of the cluster-aware validation schemes examined here and that decreased strongly with cluster count. Subject-level partitioning should be considered essential, not optional, in clinical prediction studies with repeated-measures predictors and a subject-level outcome, and cluster-aware partitioning should be applied at every level of any nested resampling procedure.

## List of Abbreviations

AR(1): first-order autoregressive process;
AUROC: area under the receiver operating characteristic curve;
CI: confidence interval;
CV: cross-validation;
DOL: day of life;
EPV: events per variable;
FiO: fraction of inspired oxygen;
ICC: intraclass correlation coefficient;
LOCO: leave-one-cluster-out;
NICU: neonatal intensive care unit;
ROP: retinopathy of prematurity;
SD: s: tandard deviation;
SpO: oxygen saturation;
TRIPOD: Transparent Reporting of a multivariable prediction model for Individual Prognosis Or Diagnosis.

## Declarations

### Ethics approval and consent to participate

The motivating clinical dataset was derived from a previously published retrospective study of oxygenation factors and retinopathy of prematurity among extremely low birth weight infants at Northside Hospital, Atlanta, Georgia [18], approved by the Northside Hospital Institutional Review Board. The present author was a co-investigator and biostatistician on the original publication and received written permission from the original study investigator to use the de-identified dataset for this secondary methodological analysis. The present analysis used only de-identified data, involved no patient contact, no new data collection, and no attempt to re-identify participants. The published report did not provide an IRB protocol number, and none was available to the author at the time of submission. This secondary analysis of de-identified data did not require separate IRB review under the Baylor College of Medicine policy.

### Consent for publication

Not applicable.

### Availability of data and materials

Appendices D and E describe the simulation and clinical-analysis code, respectively. The complete reproducible code, implemented with fixed random seeds, is available on GitHub (project home page: https://github.com/JosephHaganScD/cluster-aware-cv; release v2.1.0, commit bca7b21) and permanently archived on Zenodo (version-specific DOI https://doi.org/10.5281/zenodo.22083782) [27]. Operating system: platform independent (developed and executed under Windows 10, x86_64-w64-mingw32/x64). Programming language: R (version 4.6.1). Other requirements: R packages glmnet (version 5.0) and pROC (version 1.19.0.1) for the analysis scripts, and ggplot2, dplyr, patchwork, and scales for the figure script. License: MIT License. The motivating clinical dataset is not publicly available because it consists of de-identified patient-level data used with written permission from the original principal investigator and data custodian, and cannot be shared without authorization from the original investigator and any applicable institutional approvals. The per-dataset simulation results (main grid and sensitivity arm) are included in the archived Zenodo release; the simulated datasets can also be regenerated using the publicly available code.

### Authors’ contributions

JLH conceived the study, designed and performed all analyses, interpreted the results, and wrote and revised the manuscript. JLH read and approved the final manuscript.

## Data Availability

The simulation code and clinical analysis code are publicly available on GitHub (https://github.com/JosephHaganScD/cluster-aware-cv) and are permanently archived on Zenodo (https://doi.org/10.5281/zenodo.20222283). The motivating clinical dataset is not publicly available as it consists of de-identified patient-level data from a previously published retrospective study (Srivatsa et al., 2022; J Pediatr) and was used with written permission from the original principal investigator. The simulated datasets can be reproduced in full using the publicly available code.

https://doi.org/10.5281/zenodo.22083782

## Acknowledgements.

The author thanks Bharath Srivatsa, MD (Neonatology Associates of Atlanta, Northside Hospital, Pediatrix Medical Group) for granting permission to use the motivating dataset from the previously published study of oxygenation factors and retinopathy of prematurity [18].

## Appendices

### **Appendix A.** Correlation matrix for the seven oxygenation predictors in the motivating dataset (N = 101 subjects, 5615 daily observations).

| Predictor | FiO<sub>2</sub> | SpO<sub>2</sub> | Sev.<br>hyp. | Amb.<br>hyp. | Iatr.<br>hyp. | Swings | Tit.<br>idx. |
| --- | --- | --- | --- | --- | --- | --- | --- |
| Avg_FiO<sub>2</sub> | 1.000 | -0.492 | 0.270 | -0.385 | -0.002 | -0.023 | 0.213 |
| Avg_SpO<sub>2</sub> | -0.492 | 1.000 | -0.659 | 0.712 | 0.002 | -0.265 | -0.333 |
| Severe hypoxemia | 0.270 | -0.659 | 1.000 | -0.210 | 0.307 | 0.556 | 0.410 |
| Ambient<br>hyperoxemia | -0.385 | 0.712 | -0.210 | 1.000 | -0.315 | -0.214 | -0.347 |
| **Iatrogenic hyperoxemia** | -0.002 | 0.002 | 0.307 | -0.315 | 1.000 | 0.627 | 0.427 |
| **Swings** | -0.023 | -0.265 | 0.556 | -0.214 | 0.627 | 1.000 | 0.395 |
| **Titration index** | 0.213 | -0.333 | 0.410 | -0.347 | 0.427 | 0.395 | 1.000 |

*Pearson correlations among daily observation-level values (5615 observations, 101* *subjects). Column abbreviations: $FiO_2$ , $Avg\_FiO_2$ ; $SpO_2$ , $Avg\_SpO_2$ ; Sev. hyp.,* *severe hypoxemia; Amb. hyp., ambient hyperoxemia; Iatr. hyp., iatrogenic hyperoxemia;* *Tit. idx., titration index. Maximum absolute off-diagonal correlation = 0.712 ( $Avg\_SpO_2$* *and ambient hyperoxemia).*

### **Appendix B.** Within-cluster dependence structure (ICC and median lag-1 autocorrelation with IQR) for the seven oxygenation predictors.

| **Predictor** | **Mean** | **SD** | **ICC** | **Lag-1**<br>**median** | **Lag-1**<br>**Q25** | **Lag-1**<br>**Q75** |
| --- | --- | --- | --- | --- | --- | --- |
| **Avg_ $FiO_2$** | 32.68 | 12.29 | 0.528 | 0.727 | 0.628 | 0.832 |
| **Avg_ $SpO_2$** | 91.50 | 1.77 | 0.302 | 0.568 | 0.386 | 0.739 |
| **Severe hypoxemia** | 0.04 | 0.04 | 0.172 | 0.602 | 0.479 | 0.708 |
| **Ambient hyperoxemia** | 0.08 | 0.16 | 0.384 | 0.584 | 0.388 | 0.779 |
| **Iatrogenic hyperoxemia** | 0.14 | 0.10 | 0.163 | 0.452 | 0.334 | 0.582 |
| **Swings** | 26.00 | 29.36 | 0.189 | 0.637 | 0.512 | 0.726 |
| **Titration index** | 1.30 | 1.00 | 0.324 | 0.482 | 0.347 | 0.617 |

Mean and SD are computed across all 5615 daily observations. ICC is the one-way random-effects intraclass correlation coefficient (ICC(1,1) of Shrout and Fleiss [19]); lag-1 autocorrelation is the median (with 25th and 75th percentiles) of within-subject lag-1 autocorrelations, subjects ordered by day of life.

**Appendix C.** Covariance structure and the interpretation of the ICC by autocorrelation interaction.

For a single predictor generated under the model of Section 2.7, x_it = b_i + e_it, where b_i ∼ N(0, ICC) is the cluster-level random effect and e_it follows the stationary AR(1) process e_it = ρe_{i,t−1} + ε_it with innovation variance σ²_ε = (1 − ICC)(1 − ρ²), the marginal variance of e_it is 1 − ICC and the total marginal variance of x_it is ICC + (1 − ICC) = 1, matching the unit total variance imposed in Section 2.7. Because b_i is shared across all t and e_it is independent of b_i, the covariance between two observations from the same cluster at times j and k is Cov(x_ij, x_ik) = Var(b_i) + Cov(e_ij, e_ik) = ICC + (1 − ICC)ρ^|j−k|, so that the within-cluster correlation at lag |j − k| decomposes additively into a constant term contributed by the between-subject random effect (ICC) and a term that decays geometrically with lag through the within-subject AR(1) process ((1 − ICC)ρ^|j−k|). At lag zero this expression correctly returns the total marginal variance of 1. As the lag grows large, the correlation does not decay to zero but converges to ICC, the asymptote set by the shared random effect. This is the formal counterpart to the informal distinction drawn in Section 1 between indirect leakage through the outcome (mediated by ICC, a property of the entire cluster) and direct leakage through temporal proximity (mediated by ρ, a property that weakens with lag).

This decomposition clarifies why naive observation-level CV is optimistic through two channels rather than one. When a subject contributes observations to both the training and test folds, the test-fold predictions benefit from the shared random effect b_i regardless of which time points were observed in training (the ICC channel), and additionally benefit from the AR(1) correlation between nearby time points when the training and test observations for that subject happen to be temporally close (the ρ channel). The two channels are not independent contributors to leakage. They operate on the same underlying quantity, Cov(x_ij, x_ik), and their relative contributions shift with ICC as described below.

The interaction between ICC and ρ reported in Table 5 follows directly from this covariance structure. Write the within-cluster correlation as ρ(|j − k|) = ICC + (1 − ICC)ρ^|j−k|, and consider its sensitivity to ρ at fixed lag: ∂ρ(|j−k|)/∂ρ = (1 − ICC)·|j−k|·ρ^(|j−k|−1). This derivative is proportional to (1 − ICC), so the marginal contribution of the autocorrelation parameter to within-cluster correlation shrinks as ICC increases, vanishing entirely in the limiting case ICC = 1 (a time-invariant covariate, for which ρ is not identified because e_it degenerates to zero). This is the formal counterpart to the empirical pattern in Table 5: at ICC = 0.1, increasing ρ from 0.2 to 0.8 raised the partitioning component from +0.115 to +0.139, a 20% relative increase, whereas at ICC = 0.5 the same increase in ρ raised the partitioning component only from +0.173 to +0.178, a 3% relative increase. The (1 − ICC) weight on the autocorrelation term is largest when ICC is small, so ρ has the most room to move the within-cluster correlation, and therefore the most room to inflate naive-CV optimism, precisely when the between-subject channel is weak. Conversely, when ICC is large, the constant ICC term already places a high floor under the within-cluster correlation at every lag, so additional autocorrelation has little further leverage; clusters are already easy to identify from any single observation, independent of temporal proximity to the training data.

This qualitative account is necessarily approximate for AUROC, since AUROC is a nonlinear functional of the fitted model’s predicted probabilities rather than of the predictor covariance directly, and the fitted ridge model aggregates the (1 − ICC)ρ^|j−k| structure across eight correlated predictors rather than one. The (1 − ICC) attenuation of the ρ effect nonetheless provides a parsimonious explanation for why the ICC-by- autocorrelation interaction in Table 5 takes the sign and approximate shape that it does, and for why the N-by-ICC attenuation visible in Figure 4 behaves analogously: at small N the between-subject random effects b_i are estimated with more relative noise, so the AR(1) channel retains more influence, whereas at large N the b_i are estimated precisely enough that the ICC channel alone accounts for most of the separation between clusters, again diminishing the marginal contribution of ρ. A full analytical treatment relating this covariance decomposition to the fitted AUROC, rather than to the underlying predictor correlation, is beyond the scope of the present study and is treated qualitatively throughout the main text (Sections 1 and 4.2).

### Appendix D.

Simulation study code (R): simulation_factorial_v5.R, including the full factorial runner, the signal-variance-controlled sensitivity arm, the true-performance reference computation, results aggregation, and the reproducibility artifact (run metadata, per-condition logs, md5 checksums, sessionInfo), implemented with fixed random seeds. The main grid (162 conditions, 81,000 datasets) and the sensitivity arm (24 conditions, 24,000 dataset-arm rows) were executed under R 4.6.1 with glmnet 5.0 and pROC 1.19.0.1. Because generation was conditioned on at least three events and three non-events, the realized subject-level prevalence exceeded the nominal target in the smallest, lowest-prevalence cells; the mean realized prevalence (nominal in parentheses) was, at N = 20, 0.18 (0.10), 0.23 (0.20), and 0.35 (0.35), and at N = 30, 0.14 (0.10), 0.21 (0.20), and 0.35 (0.35), whereas for all conditions with N ≥ 50 the realized and nominal rates agreed to within 0.01. These values are means over the 500 datasets per condition, with the realized rate recorded per dataset (actual_event_rate) in the deposited simulation output. This simulation script corresponds to GitHub release v2.1.0 (commit bca7b21), archived on Zenodo at https://doi.org/10.5281/zenodo.22083782.

### Appendix E.

Clinical illustration analysis code (R): clinical_analysis_v5.R, a single script producing the cross-validation comparison, the subject-level summary models, the fold-wise coefficient stability, the regularization sensitivity analysis, and the subject-level bootstrap confidence interval for the LOCO AUROC. All clinical results are computed under one fitting convention, so that the LOCO point estimate and its confidence interval derive from the same set of model fits.

